# Prediabetes Blunts CD26/DPP4 Genetic Control of Postprandial Glycemia and Insulin Secretion

**DOI:** 10.1101/2021.07.24.21261077

**Authors:** Rita S. Patarrão, Nádia Duarte, Rogério T. Ribeiro, Rita Andrade, João Costa, Isabel Correia, José Manuel Boavida, Rui Duarte, Luís Gardete-Correia, José Luís Medina, João F. Raposo, M. Paula Macedo, Carlos Penha-Gonçalves

**Affiliations:** Centro de Estudos de Doenças Crónicas (CEDOC), NOVA Medical School-FCM, Universidade Nova de Lisboa, Lisboa, Portugal; Instituto Gulbenkian de Ciência, Rua da Quinta Grande, 6, Oeiras, Portugal; Sociedade Portuguesa de Diabetologia, Lisboa, Portugal; APDP-Diabetes Portugal Education and Research Center (APDP-ERC), Lisboa, Portugal; Departamento de Ciências Médicas, Instituto de Biomedicina - iBiMED, Universidade de Aveiro, Aveiro, Portugal

**Keywords:** CD26/DPP4, Dysglycemia, Genetic association, Highcaloric diet, Hyperinsulinemia, Insulin secretion, Postprandial glycemia, Prediabetes

## Abstract

**Aims/hypothesis:** Imbalances in glucose metabolism are hallmarks of clinically silent prediabetes representing dysmetabolism trajectories leading to type 2 diabetes (T2D). CD26/DPP4 is a clinically proven molecular target of diabetes-controlling drugs but the CD26 gene control of disglycemia is unsettled.

**Methods:** We dissected the genetic control of postprandial glycemic and insulin release responses by the CD26/DPP4 gene in an European/Portuguese population-based cohort that comprised individuals with normoglycemia and prediabetes, and in mouse **experimental models of CD26 deficiency and hypercaloric diet**.

**Results:** In individuals with normoglycemia, CD26/DPP4 single-nucleotide variants governed glycemic excursions (rs4664446, P=1.63×10^−7^) and C-peptide release responses (rs2300757, P=6.86×10^−5^) upon OGTT. Association with glycemia was stronger at 30min OGTT but the higher association of the genetic control of insulin secretion was detected in later phases of the postprandial response suggesting that CD26/DDP4 gene directly senses glucose challenges. Accordingly, in mice fed normal chow diet but not high fat diet, we found that under OGTT expression of CD26/DPP4 gene is strongly down-regulated at 30min in the mouse liver. Strikingly, no genetic association was found in prediabetic individuals indicating that postprandial glycemic control by CD26/DPP4 is abrogated in prediabetes. Furthermore, CD26 null mice provided concordant evidence that CD26/DPP4 modulates postprandial C-peptide release in normoglycemic but not in dysmetabolic states.

**Conclusions/interpretation:** These results unveiled CD26/DPP4 gene as a strong determinant of postprandial glycemia via glucose sensing mechanisms that are abrogated in prediabetes. We propose that impairments in CD26/DDP4 control of postprandial glucose-insulin responses are part of molecular mechanisms underlying early metabolic disturbances associated with T2D.

**RESEARCH IN CONTEXT:** *What is already known about this subject?:*

- CD26/DPP4 is a clinically proven molecular target of diabetes-controlling drugs
- DPP4 enzymatic activity impacts in glycemia by degrading incretins
- DPP4 represents a potential link between dysmetabolism and insulin resistance.

*What is the key question?:*

- Is CD26/DPP4 controlling postprandial glycemic and pancreatic responses?

*What are the new findings?:*

- Functional link between postprandial glycemia excursions and subsequent insulin release is operational in NGT individuals but is altered in prediabetes.
- CD26/DPP4 gene is a robust component of mechanisms that control postprandial glycemia excursions and insulin secretion in NGT individuals.
- CD26/DPP4 gene controls postprandial glucose-insulin responses via glucose sensing mechanisms that are abrogated in prediabetes.
- Postprandial reduction of hepatic CD26/DPP4 gene expression is abrogated in experimental diet-induced dysglycemia.
- CD26/DPP4 gene action in reducing post-prandial insulin release is not operating in experimental diet-induced dysglycemia.

*How might this impact on clinical practice in the foreseeable future?:*

- Impairments in the control of postprandial glucose-insulin responses by CD26/DDP4 are indicators of early metabolic disturbances and possible predictors of dysmetabolic trajectories leading to T2D.

## Introduction

Responses to blood glucose variations are afforded by the concurrent action of distinct mechanisms that maintain glycemia homeostasis in healthy individuals [1–3]. Inefficient glycemia homeostasis underlies clinically-silent dysmetabolic states representing early stages of development of type 2 diabetes (T2D). Cross-sectional studies consistently find that prediabetes has high prevalence among apparently healthy individuals in many countries and it is estimated that more than 470 million people will be prediabetic by the year 2030 [4, 5]. It is widely accepted that prediabetes is a heterogeneous condition that encompasses a diverse set of dysglycemic trajectories and therefore is a poor predictor of overt T2D development [5, 6]. Similarly to T2D, the etiopathogenesis of prediabetes is multifactorial and fueled by interactions between genetic and environmental factors.

The genetics basis of T2D susceptibility has been intensively investigated and genome-wide association studies (GWAS) using multicentric approaches had identified over 120 distinct genetic loci as T2D risk factors [7, 8]. Nevertheless, these genetic variants only account for approximately 20% of T2D heritability, possibly owing to genetic heterogeneity and to interactions with environmental factors [8, 9]. This implies that diverse pathogenesis and genetic mechanisms intervene in the course of T2D natural history [9] making it difficult to pinpoint early pathogenesis factors contributing to overt T2D. In this scenario genetic analysis of pre-disease states offers an opportunity to identify genetic modifiers of glycemic homeostatic control and to unveil mechanisms of glucose metabolism dysregulation preceding T2D development. It has been shown that genotypes predisposing to prediabetes are associated to β-cell function and insulin secretion [10]. Accordingly, a sound number of genes were found to be associated both with diabetes and glucose impairment traits in healthy individuals [9]. This illustrates that investigating genetic mechanisms that control insulin and glucose metabolism in normoglycemic individuals are instrumental to elucidate early metabolic impairments in diabetogenesis [9–11].

Dipeptidyl-Peptidase-4 (DPP4/CD26) is a ubiquitous glycoprotein occurring in two isoforms, a cell membrane-bound protein expressed in the surface of many cell types and a soluble form found in most body fluids [12, 13]. DPP4/CD26 exhibits enzymatic activity targeting a variety of substrates including cytokines, chemokines, neuropeptides and growth factors, but most prominently two incretin hormones: glucagon-like peptide-1 (GLP-1) and glucose-dependent insulinotropic polypeptide (GIP, gastric inhibitory polypeptide). GLP-1 and GIP are gut hormones released in response to digestion and absorption of food in the small intestine that act as incretins inducing postprandial insulin secretion. Incretin degrading activity of DPP4 represent a potential link between dysmetabolism and insulin resistance [14]. Serum amounts and activity levels of DPP4 are altered in many pathophysiological conditions, such as cancers, allergic asthma, hepatitis C, hepatotoxic damage, neurological and inflammatory diseases as well as in obesity and diabetes [14–18].

It is well established that DPP4 enzymatic activity impacts in glycemia by degrading incretins thereby cancelling their insulinotropic effect and leading to higher blood glucose levels. It has been shown that exposure to increased glucose levels down-regulate CD26/DPP4 expression in several cellular systems [19, 20] suggesting a glucose sensing mechanism that potentially limits DPP4 anti-incretin activity with subsequent blood glucose lowering effects. Thus, DPP4 plays intricate physiological roles in glycemic control and is unknown if postprandial glycemia responses are controlled by CD26/DPP4.

The impact of DPP4 in glucose metabolism is demonstrated by the extensive clinical usage of DPP4 inhibitors to improve glycemic control by limiting the rapid degradation of GLP-1 [21]. Further, it has been shown that the incretin effect (increased insulin secretion attributable to incretins) is markedly reduced in patients with T2D, which is believed to contribute to the impairments in glucose tolerance after food ingestion [22, 23]. Likewise, it has been shown that GLP-1 levels are reduced in prediabetes [24, 25]. This indicates that the postprandial glucose/insulin axis is impaired in early dysmetabolic stages but the involvement of CD26/DPP4 remains to be determined.

Genetic ablation of *Dpp4* in mice [26] improves insulin sensitivity and liver glucose metabolism while pharmacological inhibition of DPP4 reduces development of hepatic steatosis and improves insulin sensitivity in mouse models of obesity [27] and diabetes [28]. It was demonstrated that diet-induced obesity in mice is associated with increased expression and release of hepatic DPP4 that in turn correlates with early insulin resistance and development of liver steatosis [29]. Yet, it remains unknown whether *Dpp4* plays a biologically relevant role in glucose and insulin metabolism during the initial steps of metabolic dysregulation. We addressed these questions by investigating the genetic control of CD26/DPP4 in postprandial glycemic responses and insulin secretion in normoglycemic and prediabetic individuals and in mouse models of diet induced dysglycemia.

## Material and Methods

### Ethics statement

All subjects were volunteers and provided written informed consent for participation in this study. Ethical permits to conduct this study were obtained from the Ethics Committee of the Associação Protectora dos Diabéticos de Portugal (APDP) and from the Instituto Gulbenkian de Ciência (IGC) Ethics Committee. The study protocol adhered to the Declaration of Helsinki and was approved by the Autoridade Nacional de Protecção de Dados (permit nr._3228/2013). All procedures involving laboratory mice were in accordance with national (Portaria_1005/92) and European regulations (European Directive_86/609/CEE) on animal experimentation and were approved by the IGC Ethics Committee and the Direcção-Geral de Alimentação e Veterinária, the national authority for animal welfare.

### Subjects

The study population comprises the participants of a diabetes prevalence study performed in Portugal (PREVADIAB-2). The PREVADIAB-1 study recruited 5,167 subjects attending the national health care system across the country that were screened for diabetes status between 2008 and 2009 [30]. In 2014, we randomly selected within this cohort 1,084 non-diabetic subjects attending 55 health units geographically spread in continental Portugal. For all participants, a letter was sent containing information for study participation. For each participant medical history was assessed, BMI was recorded and routine blood tests were performed.

### Inclusion Criteria

Participants underwent a standardized 75g oral glucose tolerance test (OGTT). Venous blood samples (12hours fasting) were drawn at baseline (0min) and at time-points 30 and 120min of the OGTT. The diabetes status of each participant was determined using the World Health Organization criteria for increased risk of diabetes and prediabetes (WHO 2016, Global Report on Diabetes). Participants fulfilling the criteria for diabetes (fasting_glucose≥126mg/dl, 2h-postprandial_glucose≥200mg/dl) were excluded from this study. Participants fulfilling the criteria for impaired fasting glucose-IFG (fasting_glucose:110-125mg/dl) and/or impaired glucose tolerance-IGT (2h-postprandial_glucose:140-200mg/dl) were classified as prediabetic subjects (n=233). The remaining subjects were classified as normal glucose tolerant (NGT, n=736).

### Biochemical parameters

Glucose, insulin and C-peptide levels were measured at baseline (0min), 30 and 120min of OGTT. Plasma glucose levels were measured with a glucose analyzer (Olympus AU640, Beckman Coulter). Plasma insulin and C-peptide levels were determined by commercial chemiluminescence assays, Liaison (DiaSorin, Italy). Normal glucose tolerant (NGT) subjects and prediabetic subjects were compared using Mann-Whitney test in GraphPad Prism 7.0. The HOMA-IR [31] was assessed. For NGT versus prediabetes genetic analysis significantly different glucose and C-peptide parameters were used to estimate excursions during the OGTT according to the trapezoid method:

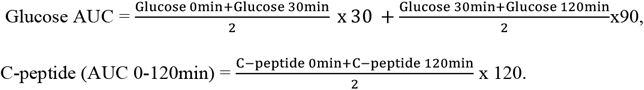

### Genotyping

Genomic DNA was extracted from whole blood using the Chemagen Magnetic Beads technology. DNA preparations were quantified using PicoGreen reagents (Invitrogen, Portugal) according to the supplier instructions. A total of 38 single nucleotide polymorphisms (SNPs) covering the *CD26* region in chromosome 2 (161.9-162.1Mb) were genotyped using the Sequenom iPlex assay (San Diego, USA) and the Sequenom MassArray K2 platform at the Genomics Unit of the IGC. Genotyping calling quality was controlled using two samples with known genotypes. Genotype calling were performed blinded to affection status. SNPs deviating from Hardy-Weinberg equilibrium (HWE) were excluded (P<0.05). The final dataset under analysis comprised 969 subjects with non-missing genotyping rate above 95%, genotyped for 33 SNPs that passed the exclusion criteria (minor allele frequency < 8% or call rate < 99%) (Supplementary_Table_1). Linkage disequilibrium (LD) map was generated by Haploview 4.2 software (Supplementary_Table_1).

### Genetic association analysis

Quantitative trait association analysis was performed using the PLINK software package, and nominal P-values for 33 CD26 SNPs were obtained for allelic and genotypic association under the additive model using age and BMI (Body Mass Index) as covariates. Empirical P values were obtained using the max(T) permutation procedure implemented in the PLINK package with 1×10^6^ label permutations Data storage and interfacing with PLINK package used the BC|Gene platform (version 3.6-036).

### Mouse Studies

C57BL/6-DPP4tm1Nwa/Orl (CD26ko) mice were purchased from European Mouse Mutant Archive (Infrafrontier Gmbh, Germany). C57BL/6 mice and CD26ko mice were bred and housed under a 12-hr light/dark cycle in specific pathogen free housing facilities at the IGC. For hypercaloric regimen experiments, C57BL/6 and CD26ko female mice with 5-6 weeks of age were maintained on regular chow (chow-RM3A from special diets services, UK) or on hypercaloric diet (HCD-catalog#TD.88137, Harlan, USA) with 42% Kcal fat (61.8% from saturated fatty acids) and 42.7%Kcal carbohydrates (63% from sugar) during 6 weeks, with free access to food and water. Oral glucose tolerance test (OGTT) was performed after overnight fasting. Mice fed with regular chow or hypercaloric diet (HCD) were gavaged with glucose at 1.5g/Kg of body weight and blood samples were collected at baseline (0min) and at 15, 30, 60 and 120min post-gavage. C-peptide was quantified using mouse-C-peptide ELISA Kit (Crystal Chem Inc., USA). C-peptide AUC was calculated using the trapezoid method, as described in human studies.

For gene expression analysis C57BL/6 mice were fasted over-night and, 30 minutes after gavage with glucose at 1.5g/Kg of body weight (glucose group) or water (control group), the liver and around 2cm pieces of duodenum, jejunum and ileum from the small intestine were collected. C57BL/6 mice maintained on HCD for 12 weeks were fasted overnight and divided into a group gavaged with glucose at 1.5g/Kg of body weight (glucose group) and a group gavaged with water (control group). 30 minutes after gavage, the liver was harvested. RNA was extracted using RNeasy Mini Kit (Qiagen, Hilden, Germany) following the manufacturer instructions, converted to cDNA (First Strand cDNA Synthesis Kit, Roche) and amplified using *cd26* (Mm00494538_m1) Taqman Gene expression assays (Applied Biosystems, USA). Endogenous Gapdh control (Mouse GAPD Endogenous Control, Applied Biosystems) was used in multiplex PCR with the target gene. PCR reactions were performed with the ABI QuantStudio-384 (Applied Biosystems). Relative quantification was calculated by the 2-ΔΔCT analysis method and expression values were normalized to the average value of the control group in the liver and to the duodenum control average value for the small intestine. Blood glucose levels were measured before and 30 minutes after gavage.

## Results

### Glucose and C-peptide postprandial responses in normoglycemic and prediabetic individuals

The PREVADIAB-2 study population is a cross-sectional sample of the European/Portuguese population-based cohort attending the national public health care system that was collected in 2014. It comprises 1,084 individuals that had been evaluated as non-diabetic 5 years prior their enrolment. We evaluated postprandial glucose metabolism in PREVADIAB-2 individuals by analyzing glycemia, insulinemia and C-peptide levels in fasting, and at 30min and 120min of a standard oral glucose tolerance test (OGTT). Blood glucose levels at fasting and at 120min after glucose challenge, revealed that 24% of the 969 non-diabetic individuals fulfilled the WHO established criteria for prediabetes. As expected, fasting plasma insulin levels, insulin resistance (HOMA-IR) and BMI were higher in the prediabetic individuals as compared to the normal glucose tolerance group (NGT) (Table_1).

**Table 1.**
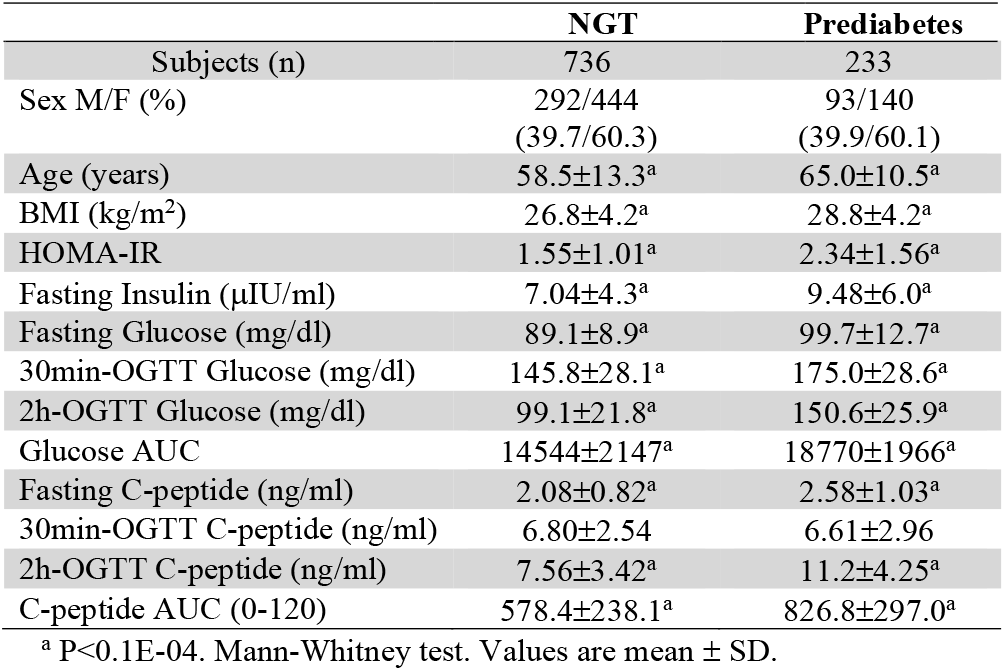
Glucose metabolism parameters of NGT and prediabetes subjects in the PREVADIAB-2 cohort.

|  | **NGT** | **Prediabetes** |
| --- | --- | --- |
| Subjects (n) | 736 | 233 |
| Sex M/F (%) | 292/444<br>(39.7/60.3) | 93/140<br>(39.9/60.1) |
| Age (years) | 58.5±13.3<sup>a</sup> | 65.0±10.5<sup>a</sup> |
| BMI (kg/m<sup>2</sup>) | 26.8±4.2<sup>a</sup> | 28.8±4.2<sup>a</sup> |
| HOMA-IR | 1.55±1.01<sup>a</sup> | 2.34±1.56<sup>a</sup> |
| Fasting Insulin (μIU/ml) | 7.04±4.3<sup>a</sup> | 9.48±6.0<sup>a</sup> |
| Fasting Glucose (mg/dl) | 89.1±8.9<sup>a</sup> | 99.7±12.7<sup>a</sup> |
| 30min-OGTT Glucose (mg/dl) | 145.8±28.1<sup>a</sup> | 175.0±28.6<sup>a</sup> |
| 2h-OGTT Glucose (mg/dl) | 99.1±21.8<sup>a</sup> | 150.6±25.9<sup>a</sup> |
| Glucose AUC | 14544±2147<sup>a</sup> | 18770±1966<sup>a</sup> |
| Fasting C-peptide (ng/ml) | 2.08±0.82<sup>a</sup> | 2.58±1.03<sup>a</sup> |
| 30min-OGTT C-peptide (ng/ml) | 6.80±2.54 | 6.61±2.96 |
| 2h-OGTT C-peptide (ng/ml) | 7.56±3.42<sup>a</sup> | 11.2±4.25<sup>a</sup> |
| C-peptide AUC (0-120) | 578.4±238.1<sup>a</sup> | 826.8±297.0<sup>a</sup> |

<sup>a</sup> P<0.1E-04. Mann-Whitney test. Values are mean ± SD.

Analysis of blood glucose levels during OGTT evaluated both by point-wise analysis or by the area under the curve (Glucose AUC) (Table_1 and Figure_1A) confirmed the expected increase of glycemic excursions in prediabetic individuals when compared to NGT [32]. Consequently, C-peptide response at the end of OGTT (120min) was clearly higher in prediabetics as compared to NGT (Table_1 and Figure_1B) supporting the notion that postprandial pancreatic insulin release is augmented in prediabetes. As expected, β-cell function as measured by the correlation between plasma glucose levels (AUC) and C-peptide levels (AUC_0-120min) was higher in NGT individuals as compared to prediabetics (Figure_1C) suggesting that the functional link between postprandial glycemia excursions and subsequent insulin release is operational in NGT individuals but somewhat affected in prediabetes.

**Figure 1.**
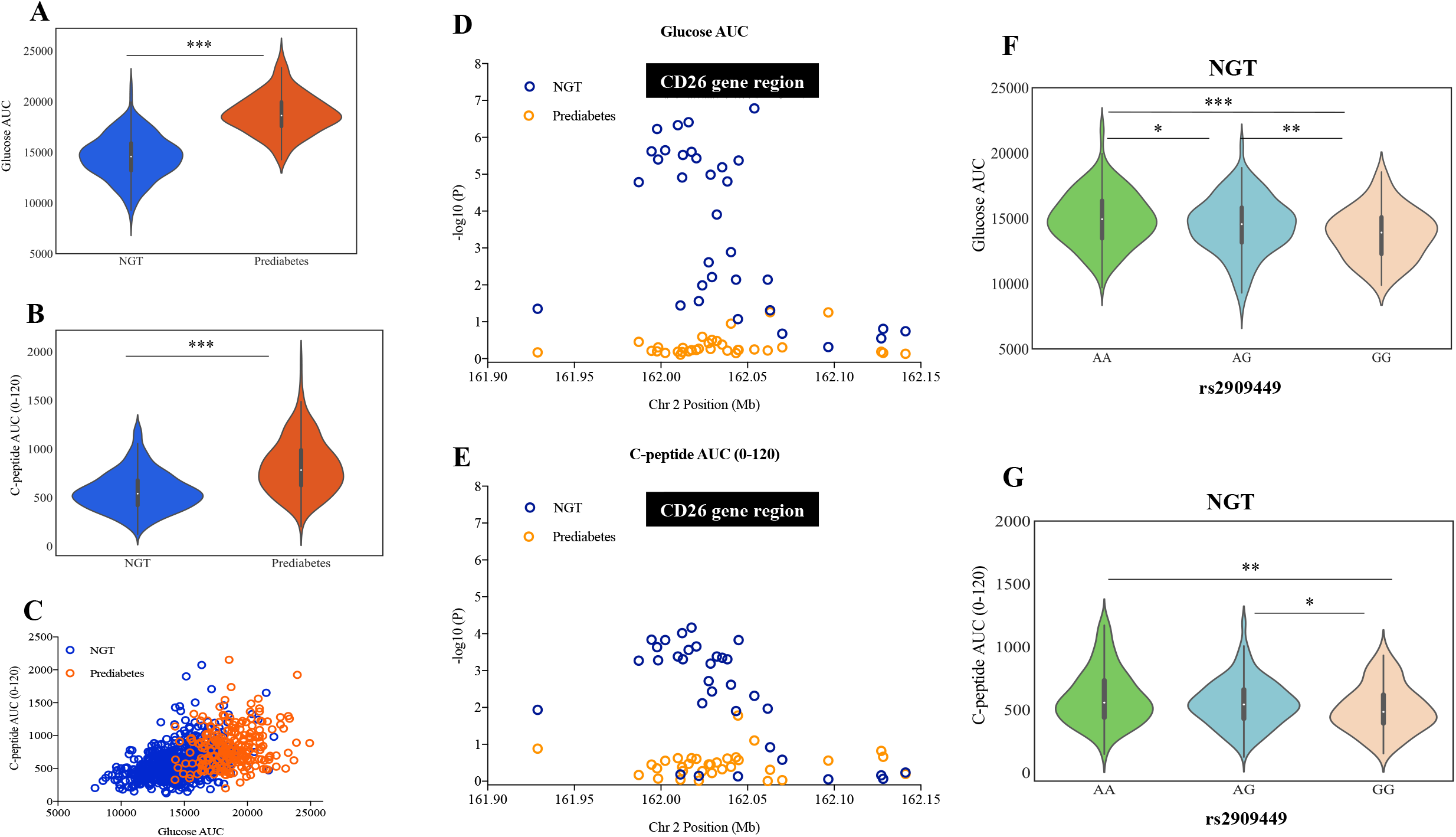
CD26/DDP4 genetic association to postprandial plasma glucose and C-peptide levels in normoglycemic (NGT) and prediabetic subjects. (A) The area under the curve (AUC) of the glucose excursion and (B) C-peptide response (AUC 0-120min) were calculated for each subject. The violin plots represent the probability density, the median, the interquartile range and the 95% confidence interval of the phenotype distributions in 736 NGT (blue) and 233 prediabetic (orange) subjects. ***P<0.001, unpaired t-test Mann-Whitney. (C) Correlation of plasma glucose AUC and C-peptide AUC (0-120) during OGTT, in NGT (blue circles; Pearson’s correlation, r^2^=0.193, P<0.0001) and in prediabetic subjects (orange circles; Pearson’s correlation, r^2^=0.055, P=0.0002). (D, E) Plots of quantitative trait locus (QTL) analysis for 33 SNPs in the CD26/DPP4 gene region testing for association with (D) plasma levels of glucose AUC and (E) C-peptide AUC (0-120) for NGT (blue circles and prediabetes subjects (orange circles). Results represent the nominal –log10 (P-value) for allelic association and SNPs position in Chromosome 2 is represented in Megabases and a scaled bar of CD26 gene region is overimposed. Violin plots of rs2909449 genotypic effects on plasma glucose AUC (F) and C-peptide AUC (0-120) (G) during OGTT in NGT subjects (green, ancestral allele homozygotes; blue heterozygotes; pink, minor allele homozygotes). Kruskal-Wallis test with Dunn’s correction for multiple comparisons (*P<0.05, **P<0.01 and ***P<0.001). The plots represent the probability density, the median, the interquartile range and the 95% confidence interval of the phenotypic distributions per genotype class totaling 736 normoglycemic subjects.

### CD26/DPP4 gene variants control glucose responses in normoglycemic subjects

We investigated whether the gene coding for CD26/DPP4 was involved in the postprandial glycemia. Non-diabetic subjects of the PREVADIAB-2 cohort (n=969) were genotyped for 33 common SNPs covering a genomic region with circa 212Kb that encompassed the CD26/DPP4 gene (Supplementary_Table_1). A large block of linkage disequilibrium (LD) was identified that spans from intron 5 to intron 25 (Supplementary_Figure_1). The role of CD26/DPP4 genetic variance in controlling glucose metabolism was separately evaluated in NGT and prediabetic individuals.

Quantitative trait locus (QTL) analysis in NGT individuals under allelic (Figure_1D) and additive models (Supplementary_Table_2) revealed highly significant association with glucose (AUC). Strong nominal association with glucose (AUC) was found across the CD26/DPP4 gene region that spans from intron 2 to intron 25. The highest association was found with rs4664446 in intron 2 (P=1.6×10^−7^, allelic model; P=1.17×10^−6^, additive model) mapping outside of the large LD block in CD26 gene. Probably representing an unlinked association signal, we found three additional peaks of association with glucose (AUC) within the LD region at rs2909449 in intron 20 (P=1.96×10^−6^, additive model), rs2268890 in intron 18 (P=2.27×10^−6^, additive model) and rs6432708 in intron 8 (P=2.87×10^−5^, additive model) (Figure_1D, Supplementary_Table_2). This genetic association was lost in individuals with prediabetes (Supplementary_Figure_2A).

### CD26/DPP4 gene variants control C-peptide responses in normoglycemic subjects

QTL analysis in NGT individuals under allelic and additive models also showed association profile of CD26/DPP4 gene with C-peptide levels during OGTT (AUC_0-120min) albeit with lower significance levels when compared to the plasma glucose levels (Figures_1E_and_1G, Supplementary_Table_3). This genetic association was lost in individuals with prediabetes (Supplementary_Figure_2B).

The statistical significance of these results resisted permutation tests (Supplementary_Tables_2_and_3) indicating that CD26/DPP4 gene is a robust component of mechanisms that control postprandial glycemia excursion and insulin secretion in normoglycemic individuals.

### CD26/DPP4 gene acts a glucose sensor in postprandial responses

Point-wise analysis of glycemia responses revealed that association signals in NGT subjects was generally more robust at 30 min OGTT as compared to 120min and was non-significant at fasting (Table_2, Supplementary_Table_4). This strongly suggests that the CD26/DPP4 gene exerts control of postprandial glucose excursions in an early time window. In contrast, control of C-peptide levels by CD26/DPP4 gene variants was not apparent at 30min OGTT but was stronger at 120min (Table_2, Supplementary_Table_5). Interestingly, peaks of association with glucose levels at 30min were in close proximity with the stronger association signals with C-peptide levels at 120mim raising the possibility that control of glycemia levels by CD26/DPP4 gene variants indirectly condition subsequent C-peptide secretion responses. Accordingly, analysis phenotypic effects of SNPs with highest association by genotype class revealed that alleles conferring lower glucose excursions in NGT individuals were also associated with lower C-peptide responses, as exemplified for rs2909449 in Figure_1F and 1G. Together these observations suggest that the CD26/DPP4 gene region senses postprandial glucose bolus and directly controls glucose excursions with indirect effects on insulin secretion.

**Table 2.** CD26/DPP4 association peaks with plasma glucose and C-peptide levels at 0min, 30min and 120min during the OGTT in NGT subjects.

| SNP ID | A | Glucose 0min |  | Glucose 30min |  | Glucose 120min |  |
| --- | --- | --- | --- | --- | --- | --- | --- |
| | | BETA | $P_{asympt}$ | BETA | $P_{asympt}$ | BETA | $P_{asympt}$ |
| rs2909449 | G | -0.75 | 1.02E-01 | -5.75 | **7.47E-05** | -3.38 | 2.84E-03 |
| rs2268890 | A | -0.68 | 1.37E-01 | -5.63 | **1.04E-04** | -3.49 | 2.06E-03 |
| rs6432708 | C | -0.58 | 2.17E-01 | -4.98 | **7.69E-04** | -3.28 | 4.44E-03 |
| rs4664446 | G | -1.48 | 1.10E-03 | -6.10 | **1.89E-05** | -2.62 | 1.91E-02 |

| SNP ID | A | C-peptide 0min |  | C-peptide 30min |  | C-peptide 120min |  |
| --- | --- | --- | --- | --- | --- | --- | --- |
| | | BETA | $P_{asympt}$ | BETA | $P_{asympt}$ | BETA | $P_{asympt}$ |
| rs7565794 | C | 0.11 | 1.02E-02 | 0.01 | 9.16E-01 | 0.66 | **4.04E-04** |
| rs1014444 | G | 0.12 | 4.25E-03 | 0.08 | 5.54E-01 | 0.66 | **2.86E-04** |
| rs2300757 | G | 0.12 | 5.40E-03 | 0.03 | 8.26E-01 | 0.68 | **2.91E-04** |
| rs16822665 | T | 0.12 | 7.71E-03 | 0.08 | 5.69E-01 | 0.65 | **7.03E-04** |

Abbreviations; A, minor allele; BETA, regression coefficient; $P_{asympt}$ , asymptotic P-value for linear regression under the additive model adjusted for age and BMI.

To experimentally test whether the *Dpp4* gene expression is controlled by postprandial glucose sensing we analyzed mouse *Dpp4* RNA levels upon glucose challenge. Glucose was administered orally to C57BL/6 mice and after 30 minutes blood glucose levels and *Dpp4* gene expression in liver and small intestine were analyzed (Figure_2B_and_2C). We observed the expected rise in blood glucose levels 30 minutes after glucose bolus in mice under normal diet (Figure_2A). As reported, expression of DPP4 in the small intestine was particularly high in ileum [33] but was not significantly altered with glucose ingestion in any of the intestine regions analyzed (Figure_2B). Dpp4 gene expression was significantly down-regulated in the liver of mice under normal diet that received glucose (Figure_2C). These results indicate that the liver responds to glucose challenge by decreasing expression of DPP4 [34] raising the possibility that in normoglycemic individuals CD26/DPP4 gene expression is under the control of an hepatic glucose sensor resulting in reduced postprandial glycemic excursions.

**Figure 2.**
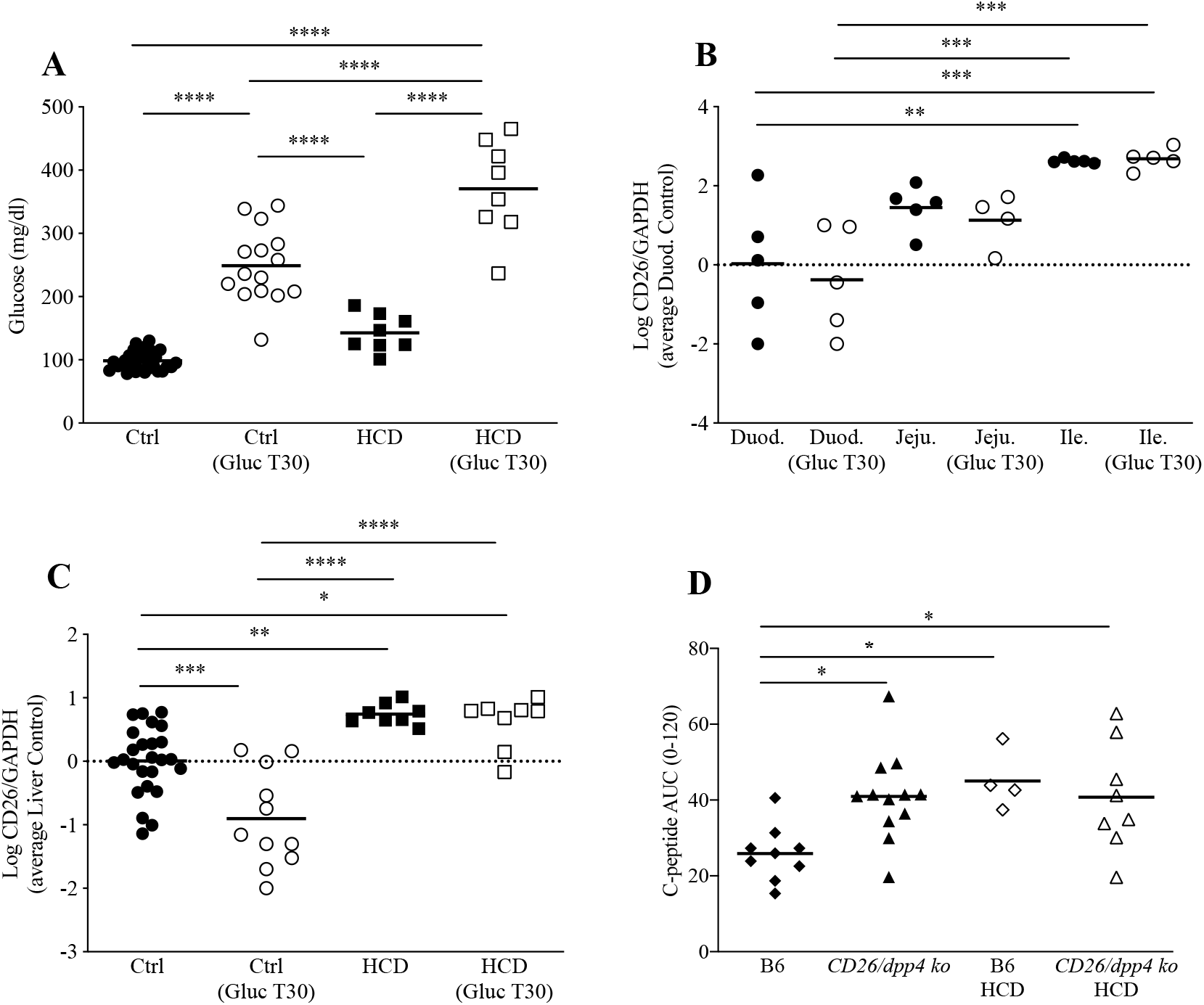
CD26/DPP4 is implicated in mouse experimental postprandial responses under normal or hypercaloric diet. Blood glucose levels before and at 30min (Gluc T30) after gavage with 1.5g/kg glucose (Gluc) (A). mRNA expression of *dpp4* (mouse gene coding for CD26/DPP4) at T30 after gavage with 1.5g/kg glucose (Gluc T30) or with water was quantified in portions of the duodenum (Duod.), jejunum (Jeju.) and ileum (Ile.) (B) as well as in the liver (C) by Real-Time PCR. Expression levels were normalized to mouse endogenous control Glyceraldehyde 3-phosphate dehydrogenase (Gapdh) and to the average value of duodenum (B) or liver (C) from control mice gavaged with water. In (B) and (C) the values are represented in a logarithmic scale. n=5-25 mice per group. Serum C-peptide levels were measured by ELISA at time 0, 15, 30, 60 and 120min during Oral Glucose Tolerance Test (OGTT) in C57BL/6 (B6) and CD26ko mice on regular chow or after 6 weeks of hypercaloric diet (HCD). The plot represents the area under the curve (AUC) from 0 to 120min (D). n=4-12 mice per group. *P<0.05, **P<0.01, ***P<0.001, ****P<0.0001. One-way ANOVA using Tukey’s correction.

### Prediabetes abrogates CD26/DPP4 genetic control of glucose and C-peptide responses

In strike contrast with normoglycemic individuals, no significant association of glucose (AUC) or C-peptide (AUC_0-120min) was found with the tested CD26/DPP4 SNPs in prediabetic subjects (Figures_1D_and_1E, Supplementary_Figure_2A_and_2B, Supplementary_Tables_2_and_3). Similarly, point-wise analysis of postprandial glycemia and C-peptide levels did not yield significant results (data not shown). These results indicate that the control of postprandial glucose mediated by the CD26/DDP4 gene is abolished in prediabetes stages possibly being implicated in early manifestations of dysglycemia.

We used a mouse model under hypercaloric diet (HCD) to mimic prediabetes and determine whether hyperglycemia affects *Dpp4* gene expression (Figure_2A_and_2C). In accordance, the glucose excursion 30 minutes after oral glucose administration was significantly higher in mice under HCD (Figure_2A). In addition, the hepatic *Dpp4* gene expression levels were increased when compared to mice on normal chow diet (Figure_2C), as previously described [29]. Surprisingly, the glucose oral bolus failed to significantly reduce liver *Dpp4* expression, as observed in normoglycemic mice, suggesting that the hepatic sensing of oral glucose is abrogated in hyperglycemic states (Figure_2C).

We further tested whether CD26/DPP4 effects on postprandial insulin secretion was impaired in dysmetabolic conditions using a mouse *Dpp4* gene knockout (ko) model. Genetic ablation of *Dpp4* allowed increased insulin secretion response during the OGTT in mice under normal chow diet confirming the notion that CD26/DPP4 acts to reduce postprandial insulin response to glucose challenge under standard metabolic conditions. Notably, this differential response was blunted in mice exposed to HCD as *Dpp4* null mice and wild-type animals showed similar increase in postprandial insulin secretion (Figure_2D). This finding corroborates our data indicating that the genetic effect of CD26/DPP4 on postprandial glycemic/insulin axis responses is abrogated in early dysmetabolic stages.

## Discussion

This study revealed the CD26/DPP4 gene controls responses to postprandial glucose challenges in normoglycemic subjects. Specifically, we found that genetic polymorphisms in the CD26/DPP4 locus, spanning from intron 2 to 25, control both glucose excursions and C-peptide release after glucose ingestion in normoglycemic but not in prediabetic individuals. Notably, genetic control of glucose excursions was more pronounced at early time points in the OGCTT while the genetic effects on insulin secretion were stronger in later phases of the postprandial response. Together the results obtained from human subjects and experimental models support the proposal of a genetic mechanism of glucose-sensing for fine-tuning the response to glucose ingestion that operates via CD26/DPP4 gene and is overridden in dysfunctional glycemic states.

Importantly, we found that CD26/DPP4 alleles acting to reduced postprandial glucose excursions were also associated to reduced C-peptide release in NGT subjects suggesting that the genetic effect on the pancreatic response was subsequent to the control over the glucose excursions (Figure_1F_and_1G, Supplementary_Table_2 and Supplementary_Table_3). In agreement, association of CD26/DPP4 SNPs with postprandial C-peptide release was not detected at 30mim but only at 120min after glucose challenge. On the other hand, association with glucose levels was readily detected at 30min (Table_2) again suggesting that the genetic control of glucose excursions conditioned the pancreatic response. This offers an explanation for the detection of stronger genetic association signals with glucose excursions (Glucose_AUC_0-120min) as compared to pancreatic responses (C-peptide_AUC_0-120min). These findings denote that control of postprandial glycemia by CD26/DPP4 governs the subsequent C-peptide release response and suggest that in normoglycemic state CD26/DPP4 gene is part of a postprandial glycemia sensing mechanism.

Genotypes at the CD26/DPP4 locus had no impact in controlling serum DPP4 enzymatic levels (not shown) suggesting that the action CD26/DPP4 SNPs in controlling postprandial glycemia and insulin secretion is not detectable by assessing DPP4 levels in peripheral blood. Several mechanisms were suggested to be involved in glucose lowering effect of DPP4 inhibitors in rodent models and in humans [21] that do not affect levels of plasma DPP4 activity. In mice, inhibition of DPP4 in the gut but not in the plasma was achieved by low dose of the DDP4 inhibitor leading to glucose regulation through local effects on GLP-1 inactivation [35]. DPP4 inhibitors were shown to prevent inactivation of GLP-1 in the gut or in pancreatic islets contributing to reduced hepatic glucose production or increased insulin secretion and decreased glucagon secretion [36]. These data suggest that CD26/DPP4 controls glycemic responses using mechanisms that do not affect DPP4 plasma levels.

Reduction of CD26/DPP4 expression upon exposure to high glucose was reported to occur *in vitro* in epithelium intestinal cell line [20] and in adipocytes [19]. We found that liver expression of CD26/DPP4 mRNA was down-regulated upon *in vivo* exposure to glucose. These results suggest that glucose-induced modulation of CD26/DPP4 expression occurs in the liver and could have an indirect impact on hepatic functions contributing to regulation of glucose excursions. Coincidentally, exogenous GLP-1 administration was shown to suppress hepatic glucose output in human subjects independently of plasma insulin, C-peptide levels and without alterations in non-esterified fatty acids levels [34]. Together, this evidence raises the possibility that the CD26/DPP4 gene senses and responds to postprandial glucose through down-regulation of RNA expression, prominently in the liver but not in the gut, possibly impacting in liver glucose output and leading to glycemic lowering effects. The specific glucose sensing mechanisms that determine the differential CD26/DPP4 gene regulation in the liver warrants further research. Several studies suggest an alternative model of postprandial GLP-1 action through a neural circuit originating in the hepatic portal area [37]. Evidence of rapid degradation of GLP-1 in the hepato-portal bed even in the presence of vildagliptin (DPP4 inhibitor) suggests that a mechanism other than endocrine action accounts for the glucose lowering of DPP4 inhibition [38]. Infusion of GLP-1 and glucose intraportally caused an increase in glucose-stimulated insulin secretion that was attenuated by neural blockade. GLP-1r is expressed by vagal afferent neurons that innervate the abdominal organs including the hepatoportal region [37, 38]. It was observed that selectively blocking the GLP-1r in the portal circulation caused a significant impairment of glucose tolerance. These findings support that vagal GLP-1r neurons innervating the hepatic portal region mediate the glucose-lowering effect of endogenous GLP-1 and that local blockade of portal GLP-1r causes glucose intolerance. These findings are consistent with a neural pathway by which GLP-1r can initiate responses from the splanchnic bed, closer to the origin of GLP-1 secretion, that promote glucose tolerance [38].

The striking finding that glucose excursion or C-peptide responses in prediabetic subjects is not controlled by CD26/DPP4 gene variants suggests that dysglycemic states abrogate the postprandial glucose sensing by CD26/DPP4. A study by Bohm et al. [39] using a cohort of overweight subjects with 70% prediabetics identified one SNP, rs6741949, in intron 2 of the CD26/DPP4 locus to be weakly associated (P=0.0447) with oral glucose-stimulated GLP-1 increase. Association of the minor allele of this SNP with increased insulin secretion was only obtain within high body fat individuals suggesting that this association with incretin secretion mainly depends on increased body adiposity [39]. Other authors also show that DPP4 genetic polymorphisms in T2D individuals are associated with serum lipid levels, where allele mutation of A to G in rs3788979 was associated with reduced ApoB level [40–42]. We also found no association of CD26/DPP4 in our group of prediabetic individuals that comprised individuals with impaired fasting glucose (IFG), with impaired glucose tolerance (IGT) and individuals with combined IFG and IGT. Nevertheless, it remains to be determined whether these prediabetes sub-phenotypes equally contribute to loss of postprandial glucose sensing by CD26/DPP4. Our finding that postprandial glucose excursions in mice exposed to HCD failed to reduce CD26/DDP4 expression reinforces the hypothesis that glucose sensing by the CD26/DDP4 gene is impaired in dysmetabolic stages. Further, this raises the possibility that epigenetic modifications imposed by exposure to sustained dysmetabolic conditions abrogate CD26/DDP4 gene action on postprandial glucose metabolism. Whether such genetic modifications would be reversable in subjects with diabetic dysmetabolism ought to be further addressed. The state of art in therapeutic recommendations only targets the overall CD26/DPP4 activity. Our animal work suggests that the lost hepatic capacity of postprandial glucose sensing by the CD26/DPP4 gene might become a relevant therapeutic driver.

In sum, our data indicate that CD26/DPP4 genetic polymorphisms take part in the control of postprandial glycemia excursions and subsequently in C-peptide release responses through glucose-sensing mechanisms that are overridden in prediabetes.

## Data Availability

The results of this paper unveiled CD26/DPP4 gene as a strong determinant of postprandial glycemia via glucose sensing mechanisms that are abrogated in prediabetes. We propose that impairments in CD26/DDP4 control of postprandial glucose-insulin responses are part of molecular mechanisms underlying early metabolic disturbances associated with T2D.

## Acknowledgments

We wish to acknowledge the participants in the PREVADIAB-2 study. The PREVADIAB-2 study was supported by a grant from the Portuguese Directorate General of Health. This work was financed by Fundação para a Ciência e Tecnologia, reference number PTDC/BIM/MET/4265/2014 and by ONEIDA, project E-411021.01 - Lisboa-01-0145-FEDER-016417 co-funded by FEEI (Fundos Europeus Estruturais e de Investimento) from Programa Operacional Regional Lisboa 2020. We also acknowledge the research infrastructure CONGENTO, project LISBOA-01-0145-FEDER-022170, co-financed by Lisboa Regional Operational Programme (Lisboa 2020), under the Portugal 2020 Partnership Agreement, through the European Regional Development Fund (ERDF) and Foundation for Science and Technology (Portugal).

## ABBREVIATIONS

APDP: Associação Protectora dos Diabéticos de Portugal
BMI: Body Mass Index
CD26: Cluster of differentiation 26
CI: Confidence Interval
DPP4: Dipeptidyl Peptidase 4
GIP: Gastric inhibitory polypeptide
GLP-1: Glucagon-like peptide-1
GWAS: Genome-Wide Association Studies
HCD: Hypercaloric Diet
HOMA-IR: Homeostatic Model Assessment
HWE: Hardy-Weinberg equilibrium
IFG: Impaired fasting glucose
IGC: Instituto Gulbenkian de Ciência
IGT: Impaired glucose tolerance
KO: Knockout
LD: Linkage disequilibrium
Mb: Megabases
NGT: Normal glucose tolerant
OGTT: Oral Glucose Tolerance Test
PREVADIAB: Diabetes prevalence study in Portugal
QTL: Quantitative trait locus
SNP: Single-Nucleotide Polymorphism
T2D: Type 2 Diabetes

---

**Supplementary Figure 1.**
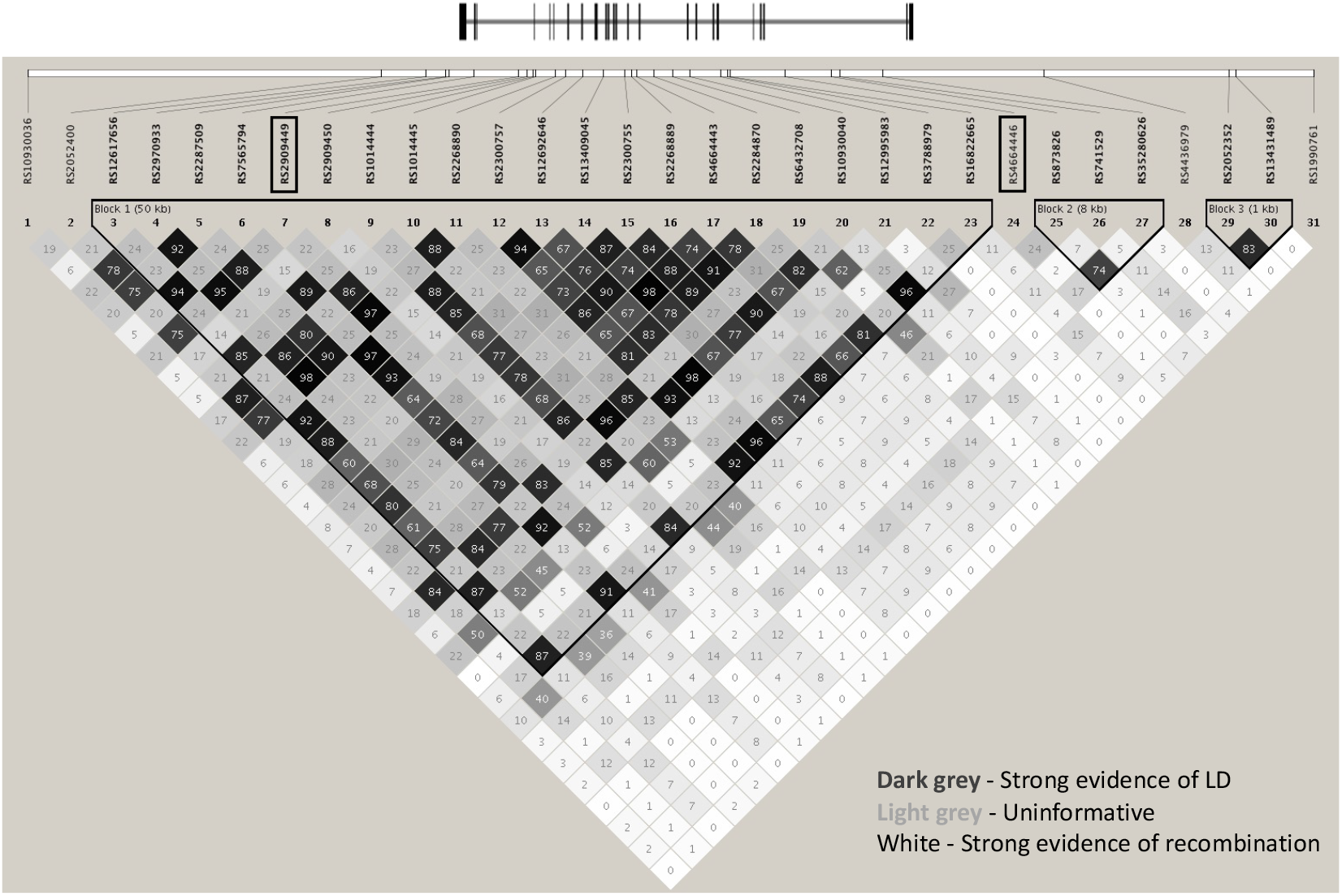
Linkage disequilibrium (LD) map of the genotyped CD26/DPP4 SNPs with overimposed scaled representation of exon-intron structure of the CD26/DPP4 gene (oriented from right to left). R-square values for pairwise LD were calculated using 969 subjects analyzed in this study. Color coding highlights a large LD block spanning from intron 5 to 25 (Block 1). Indels (rs138687963 and rs71408196) are excluded from the LD map. LD plot was generated by Haploview 4.2. software.

**Supplementary Figure 2.**
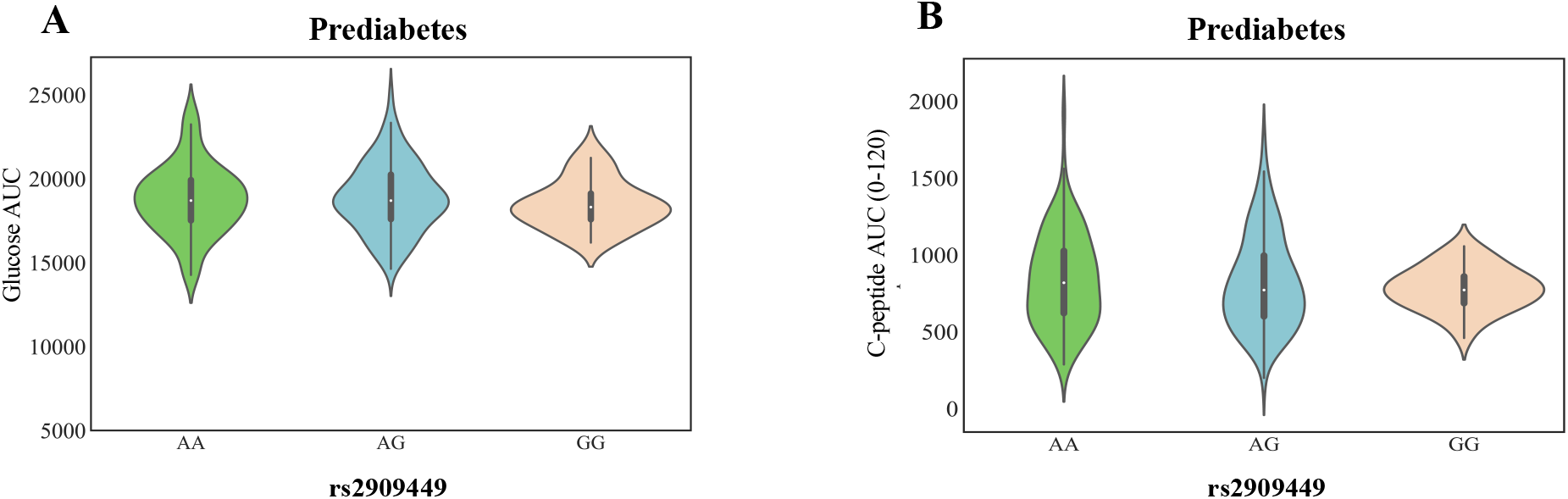
Violin plots of genotype-class effects of rs2909449 on plasma glucose AUC (A) and C-peptide AUC (0-120) (B) during OGTT (green, ancestral allele homozygotes; blue heterozygotes; pink, minor allele homozygotes). Kruskal-Wallis test with Dunn’s correction for multiple comparisons (*P<0.05, **P<0.01 and ***P<0.001, unpaired t-test Mann-Whitney). The plots represent the probability density, the median, the interquartile range and the 95% confidence interval of the phenotypic distributions per genotype class totaling 233 prediabetic subjects.

## Notes

### Competing Interest Statement

The authors have declared no competing interest.

### Funding Statement

This work was financed by Fundacao para a Ciencia e Tecnologia, reference number PTDC/BIM/MET/4265/2014 and by ONEIDA, project E-411021.01 - Lisboa-01-0145-FEDER-016417 co-funded by FEEI (Fundos Europeus Estruturais e de Investimento) from Programa Operacional Regional Lisboa 2020. We also acknowledge the research infrastructure CONGENTO, project LISBOA-01-0145-FEDER-022170, co-financed by Lisboa Regional Operational Programme (Lisboa 2020), under the Portugal 2020 Partnership Agreement, through the European Regional Development Fund (ERDF) and Foundation for Science and Technology (Portugal).

### Author Declarations

All subjects were volunteers and provided written informed consent for participation in this study. Ethical permits to conduct this study were obtained from the Ethics Committee of the Associacao Protectora dos Diabeticos de Portugal (APDP) and from the Instituto Gulbenkian de Ciencia (IGC) Ethics Committee. The study protocol adhered to the Declaration of Helsinki and was approved by the Autoridade Nacional de Protecao de Dados (permit nr._3228/2013). All procedures involving laboratory mice were in accordance with national (Portaria_1005/92) and European regulations (European Directive_86/609/CEE) on animal experimentation and were approved by the IGC Ethics Committee and the Direcao-Geral de Alimentacao e Veterinaria, the national authority for animal welfare.

