## Supplementary material for "Prediabetes Blunts CD26/DPP4 Genetic Control of Postprandial Glycemia and Insulin Secretion": Supp Material

**Supplementary Data**

**Supplementary Table 1 -** Description of 33 CD26/DDP4 SNPs genotyped in this study.

| SNP ID | bp | Alleles | MAF | Position in gene |
|---|---|---|---|---|
| rs10930036 | 161928813 | C/T | 0.33 | Downstream |
| rs2052400 | 161987158 | G/A | 0.34 | Downstream |
| rs12617656 | 161994637 | T/C | 0.29 | Intron 25 |
| rs2970933 | 161997766 | G/A | 0.38 | Intron 23 |
| rs2287509 | 161998398 | T/G | 0.37 | Intron 23 |
| rs7565794 | 162002490 | T/C | 0.29 | Intron 23 |
| rs2909449 | 162009828 | A/G | 0.38 | Intron 20 |
| rs2909450 | 162011281 | G/A | 0.28 | Intron 19 |
| rs1014444 | 162012348 | A/G | 0.31 | Intron 19 |
| rs1014445 | 162012693 | G/A | 0.35 | Intron 19 |
| rs2268890 | 162015982 | G/A | 0.37 | Intron 18 |
| rs2300757 | 162017660 | G/C | 0.28 | Intron 16 |
| rs12692646 | 162020518 | T/A | 0.28 | Intron 13 |
| rs138687963 | 162021895 | A/- | 0.28 | Intron 11 |
| rs13409045 | 162023873 | T/C | 0.36 | Intron 10 |
| rs2300755 | 162027452 | C/T | 0.33 | Intron 10 |
| rs2268889 | 162028600 | G/A | 0.30 | Intron 10 |
| rs4664443 | 162029379 | A/G | 0.36 | Intron 10 |
| rs2284870 | 162032217 | A/G | 0.32 | Intron 10 |
| rs6432708 | 162035338 | T/C | 0.35 | Intron 8 |
| rs10930040 | 162038256 | G/A | 0.28 | Intron 8 |
| rs71408196 | 162040386 | -/AT | 0.11 | Intron 5 |
| rs12995983 | 162043293 | T/C | 0.26 | Intron 5 |
| rs3788979 | 162044379 | G/A | 0.09 | Intron 5 |
| rs16822665 | 162044817 | C/T | 0.28 | Intron 5 |
| rs4664446 | 162053893 | A/G | 0.44 | Intron 2 |
| rs873826 | 162061561 | G/A | 0.32 | Intron 2 |
| rs741529 | 162062979 | G/A | 0.14 | Intron 2 |
| rs35280626 | 162070034 | C/T | 0.27 | Intron 2 |
| rs4436979 | 162096640 | T/C | 0.49 | Upstream |
| rs2052352 | 162127239 | C/T | 0.42 | Upstream |
| rs13431489 | 162128319 | A/G | 0.39 | Upstream |
| rs1990761 | 162141193 | A/G | 0.30 | Upstream |

bp (position in chromosome 2 according to GRCh38.p10); Alleles (ancestral/minor); MAF (minor allele frequency in PREVADIAB-2 cohort); Relative position in CD26 exon-intron structure. rs138687963 and rs71408196 are indels.

**Supplementary Table 2 -** Quantitative trait locus analysis of Glucose AUC during the OGTT with SNPs in the CD26/DPP4 gene region.

| | | NGT (n=736) | | | | Prediabetes (n=233) | | |
|---|---|---|---|---|---|---|---|---|
| | **A** | **BETA** | **SE** | **$P_{asymp}$** | **$P_{emp.}$** | **BETA** | **SE** | **$P_{asymp.}$** |
| rs10930036 | T | -186.20 | 113.20 | 1.01E-01 | 1.01E-01 | -45.67 | 212.50 | 8.30E-01 |
| rs2052400 | A | -433.00 | 106.70 | 5.48E-05 | 4.90E-05 | -117.80 | 207.80 | 5.71E-01 |
| rs12617656 | C | 460.60 | 114.80 | 6.69E-05 | 5.70E-05 | 8.06 | 213.90 | 9.70E-01 |
| rs2970933 | A | -494.70 | 106.50 | 4.10E-06 | 4.00E-06 | -27.87 | 197.90 | 8.88E-01 |
| rs2287509 | G | -461.20 | 108.00 | 2.23E-05 | 1.70E-05 | -97.08 | 197.50 | 6.24E-01 |
| rs7565794 | C | 463.10 | 113.90 | 5.34E-05 | 5.40E-05 | -15.99 | 212.00 | 9.40E-01 |
| **rs2909449** | **G** | **-509.00** | **106.10** | **1.96E-06** | **3.00E-06** | **-12.22** | **194.60** | **9.50E-01** |
| rs2909450 | A | 281.00 | 119.30 | 1.87E-02 | 1.89E-02 | -51.02 | 209.10 | 8.08E-01 |
| rs1014444 | G | 429.00 | 111.40 | 1.29E-04 | 1.24E-04 | 10.81 | 206.30 | 9.58E-01 |
| rs1014445 | A | -465.10 | 107.60 | 1.75E-05 | 1.90E-05 | -57.15 | 208.00 | 7.84E-01 |
| **rs2268890** | **A** | **-505.50** | **106.00** | **2.27E-06** | **3.00E-06** | **-23.69** | **197.40** | **9.05E-01** |
| rs2300757 | G | 457.10 | 115.40 | 8.29E-05 | 7.70E-05 | 22.10 | 211.90 | 9.17E-01 |
| rs12692646 | A | 468.30 | 115.80 | 5.80E-05 | 5.20E-05 | 25.04 | 215.10 | 9.07E-01 |
| rs138687963 | D | 308.30 | 119.20 | 9.91E-03 | 9.93E-03 | -129.40 | 207.40 | 5.33E-01 |
| rs13409045 | T | 209.80 | 106.80 | 5.00E-02 | 5.00E-02 | 157.00 | 201.50 | 4.37E-01 |
| rs2300755 | T | 271.30 | 109.50 | 1.34E-02 | 1.33E-02 | 99.99 | 201.00 | 6.19E-01 |
| rs22688890 | A | 409.90 | 114.60 | 3.73E-04 | 3.19E-04 | 46.39 | 204.00 | 8.20E-01 |
| rs4664443 | G | 228.30 | 106.80 | 3.29E-02 | 3.29E-02 | 128.20 | 202.60 | 5.28E-01 |
| rs2284870 | G | 346.00 | 112.30 | 2.15E-03 | 2.15E-03 | 127.60 | 201.60 | 5.27E-01 |
| **rs6432708** | **C** | **-456.40** | **108.40** | **2.87E-05** | **3.10E-05** | **-79.69** | **209.80** | **7.04E-01** |
| rs10930040 | G | 434.50 | 117.00 | 2.21E-04 | 2.06E-04 | 15.83 | 215.10 | 9.41E-01 |
| rs71408196 | I | 518.70 | 167.70 | 2.06E-03 | 2.07E-03 | 321.20 | 276.50 | 2.47E-01 |
| rs12995983 | C | -256.10 | 119.00 | 3.17E-02 | 3.18E-02 | 197.70 | 231.10 | 3.93E-01 |
| rs3788979 | A | 214.50 | 177.90 | 2.28E-01 | 2.28E-01 | -155.50 | 328.90 | 6.37E-01 |
| rs16822665 | T | 457.10 | 117.10 | 1.04E-04 | 8.80E-05 | 20.71 | 217.20 | 9.24E-01 |
| **rs4664446** | **G** | **-511.40** | **104.30** | **1.17E-06** | **3.00E-06** | **197.20** | **184.30** | **2.86E-01** |
| rs873826 | A | -260.90 | 112.90 | 2.12E-02 | 2.12E-02 | 147.30 | 208.50 | 4.81E-01 |
| rs741529 | A | 212.50 | 151.60 | 1.62E-01 | 1.62E-01 | 430.10 | 262.70 | 1.03E-01 |
| rs35280626 | T | -146.10 | 119.00 | 2.20E-01 | 2.20E-01 | 213.00 | 212.60 | 3.18E-01 |
| rs4436979 | C | -144.20 | 106.70 | 1.77E-01 | 1.77E-01 | -312.90 | 199.20 | 1.18E-01 |
| rs2052352 | T | 87.90 | 104.10 | 3.99E-01 | 3.99E-01 | 110.10 | 186.70 | 5.56E-01 |
| rs13431489 | G | 146.40 | 106.30 | 1.69E-01 | 1.69E-01 | 102.60 | 189.20 | 5.88E-01 |
| rs1990761 | G | -134.60 | 115.20 | 2.43E-01 | 2.44E-01 | 126.90 | 209.90 | 5.46E-01 |

Abbreviations: A, minor allele; BETA, regression coefficient; SE, standard error; $P_{asymp}$, asymptotic P-value for linear regression under the additive model adjusted for age and BMI; $P_{emp}$, empirical pointwise P-value ($10^6$ permutations). SNP's representing peaks of association are highlighted in bold.

**Supplementary Table 3 -** Quantitative trait loucs analysis of C-peptide AUC (0-120) during the OGTT with SNPs in the CD26/DPP4 gene region.

| | | NGT (n=736) | | | | Prediabetes (n=233) | | |
|---|---|---|---|---|---|---|---|---|
| | **A** | **BETA** | **SE** | **P*asymp*** | **P*emp.*** | **BETA** | **SE** | **P*asymp.*** |
| rs10930036 | T | -30.37 | 12.62 | 1.64E-02 | 1.65E-02 | -42.59 | 31.17 | 1.73E-01 |
| rs2052400 | A | -38.41 | 11.98 | 1.41E-03 | 1.36E-03 | -5.30 | 30.63 | 8.63E-01 |
| rs12617656 | C | 45.36 | 12.88 | 4.58E-04 | 4.58E-04 | 19.47 | 31.49 | 5.37E-01 |
| rs2970933 | A | -39.34 | 11.99 | 1.09E-03 | 1.09E-03 | -22.41 | 29.12 | 4.42E-01 |
| rs2287509 | G | -37.96 | 12.13 | 1.83E-03 | 1.79E-03 | -7.73 | 29.12 | 7.91E-01 |
| **rs7565794** | **C** | **46.21** | **12.78** | **3.21E-04** | **3.11E-04** | **21.45** | **31.20** | **4.92E-01** |
| rs2909449 | G | -38.21 | 11.92 | 1.41E-03 | 1.38E-03 | -30.21 | 28.60 | 2.92E-01 |
| rs2909450 | A | 1.14 | 13.40 | 9.33E-01 | 9.33E-01 | 14.09 | 30.80 | 6.48E-01 |
| **rs1014444** | **G** | **46.83** | **12.47** | **1.87E-04** | **1.95E-04** | **15.58** | **30.37** | **6.09E-01** |
| rs1014445 | A | -37.96 | 12.08 | 1.74E-03 | 1.72E-03 | -10.62 | 30.64 | 7.29E-01 |
| rs2268890 | A | -39.12 | 11.92 | 1.08E-03 | 1.06E-03 | -26.00 | 29.03 | 3.71E-01 |
| **rs2300757** | **G** | **48.47** | **12.97** | **2.02E-04** | **1.89E-04** | **24.01** | **31.22** | **4.43E-01** |
| rs12692646 | A | 45.54 | 13.02 | 4.98E-04 | 5.05E-04 | 27.39 | 31.64 | 3.88E-01 |
| rs138687963 | D | 1.91 | 13.41 | 8.87E-01 | 8.87E-01 | 5.54 | 30.58 | 8.56E-01 |
| rs13409045 | T | 30.87 | 11.91 | 9.78E-03 | 9.76E-03 | 4.37 | 29.73 | 8.83E-01 |
| rs2300755 | T | 37.24 | 12.22 | 2.41E-03 | 2.42E-03 | 19.83 | 29.60 | 5.04E-01 |
| rs2268889 | A | 40.22 | 12.85 | 1.83E-03 | 1.85E-03 | 20.50 | 30.03 | 4.96E-01 |
| rs4664443 | G | 33.65 | 11.92 | 4.91E-03 | 4.92E-03 | 1.18 | 29.88 | 9.69E-01 |
| rs2284870 | G | 41.46 | 12.55 | 1.01E-03 | 1.04E-03 | 24.84 | 29.67 | 4.03E-01 |
| rs6432708 | C | -38.65 | 12.16 | 1.54E-03 | 1.51E-03 | -11.84 | 30.90 | 7.02E-01 |
| rs10930040 | G | 42.46 | 13.05 | 1.19E-03 | 1.20E-03 | 28.80 | 31.63 | 3.64E-01 |
| rs71408196 | I | 55.87 | 18.79 | 3.05E-03 | 3.19E-03 | -55.13 | 40.69 | 1.77E-01 |
| rs12995983 | C | -35.04 | 13.25 | 8.36E-03 | 8.42E-03 | -26.52 | 34.05 | 4.37E-01 |
| rs3788979 | A | -8.95 | 19.96 | 6.54E-01 | 6.53E-01 | 116.80 | 47.82 | 1.54E-02 |
| **rs16822665** | **T** | **46.02** | **13.13** | **4.87E-04** | **4.99E-04** | **26.97** | **31.95** | **4.00E-01** |
| rs4664446 | G | -29.10 | 11.82 | 1.41E-02 | 1.42E-02 | -44.60 | 27.06 | 1.01E-01 |
| rs873826 | A | -28.85 | 12.58 | 2.21E-02 | 2.20E-02 | 16.92 | 30.73 | 5.82E-01 |
| rs741529 | A | 22.38 | 16.99 | 1.88E-01 | 1.88E-01 | -44.32 | 38.83 | 2.55E-01 |
| rs35280626 | T | -16.98 | 13.23 | 2.00E-01 | 2.01E-01 | 29.45 | 31.33 | 3.48E-01 |
| rs4436979 | C | -7.70 | 11.94 | 5.19E-01 | 5.20E-01 | 34.97 | 29.42 | 2.36E-01 |
| rs2052352 | T | 10.84 | 11.65 | 3.52E-01 | 3.53E-01 | -43.58 | 27.36 | 1.13E-01 |
| rs13431489 | G | 11.65 | 11.89 | 3.28E-01 | 3.28E-01 | -40.52 | 27.75 | 1.46E-01 |
| rs1990761 | G | -9.32 | 12.88 | 4.69E-01 | 4.70E-01 | -1.65 | 30.94 | 9.58E-01 |

Abbreviations: A, minor allele; BETA, regression coefficient; SE, standard error; P*asymp*, asymptotic P-value for linear regression under the additive model adjusted for age and BMI; P*emp,* empirical pointwise P-value ($10^6$ permutations). SNP's representing peaks of association are highlighted in bold.

**Supplementary Table 4 -** Quantitative trait locus analysis of plasma glucose levels at 0min, 30min and 120min during the OGTT with CD26/DPP4 SNPs in NGT subjects.

| | | | Glucose 0min | | | Glucose 30min | | | Glucose 120min | |
|---|---|---|---|---|---|---|---|---|---|---|
| **SNP ID** | **A** | **BETA** | **SE** | **$P_{asymp}$** | **BETA** | **SE** | **$P_{asymp}$** | **BETA** | **SE** | **$P_{asymp.}$** |
| rs10930036 | T | 0.24 | 0.48 | 6.19E-01 | -2.28 | 1.53 | 1.37E-01 | -1.26 | 1.19 | 2.92E-01 |
| rs2052400 | A | -0.72 | 0.46 | 1.18E-01 | -4.47 | 1.45 | 2.18E-03 | -3.42 | 1.13 | 2.58E-03 |
| rs12617656 | C | 0.98 | 0.50 | 4.85E-02 | 4.93 | 1.56 | 1.65E-03 | 3.28 | 1.22 | 7.39E-03 |
| rs2970933 | A | -0.69 | 0.46 | 1.37E-01 | -5.46 | 1.45 | 1.80E-04 | -3.49 | 1.13 | 2.15E-03 |
| rs2287509 | G | -0.63 | 0.47 | 1.81E-01 | -5.30 | 1.47 | 3.29E-04 | -2.98 | 1.15 | 9.50E-03 |
| rs7565794 | C | 0.89 | 0.49 | 7.23E-02 | 4.94 | 1.55 | 1.50E-03 | 3.36 | 1.21 | 5.66E-03 |
| **rs2909449** | **G** | -0.75 | 0.46 | **1.02E-01** | **-5.75** | **1.44** | **7.47E-05** | **-3.38** | **1.13** | **2.84E-03** |
| rs2909450 | A | 0.36 | 0.51 | 4.83E-01 | 3.55 | 1.62 | 2.82E-02 | 1.35 | 1.26 | 2.87E-01 |
| rs1014444 | G | 0.85 | 0.48 | 7.72E-02 | 4.47 | 1.51 | 3.21E-03 | 3.23 | 1.18 | 6.38E-03 |
| rs1014445 | A | -0.53 | 0.47 | 2.56E-01 | -5.07 | 1.46 | 5.59E-04 | -3.37 | 1.14 | 3.28E-03 |
| **rs2268890** | **A** | -0.68 | 0.46 | **1.37E-01** | **-5.63** | **1.44** | **1.04E-04** | **-3.49** | **1.13** | **2.06E-03** |
| rs2300757 | G | 0.92 | 0.50 | 6.67E-02 | 4.93 | 1.57 | 1.80E-03 | 3.23 | 1.23 | 8.69E-03 |
| rs12692646 | A | 0.97 | 0.50 | 5.29E-02 | 5.52 | 1.57 | 4.68E-04 | 2.67 | 1.24 | 3.13E-02 |
| rs138687963 | D | 0.37 | 0.51 | 4.74E-01 | 3.74 | 1.62 | 2.09E-02 | 1.70 | 1.26 | 1.79E-01 |
| rs13409045 | T | 0.22 | 0.46 | 6.26E-01 | 2.11 | 1.45 | 1.46E-01 | 1.80 | 1.13 | 1.11E-01 |
| rs2300755 | T | 0.46 | 0.47 | 3.33E-01 | 3.03 | 1.49 | 4.14E-02 | 1.87 | 1.16 | 1.07E-01 |
| rs2268889 | A | 0.71 | 0.49 | 1.53E-01 | 4.82 | 1.56 | 2.01E-03 | 2.39 | 1.22 | 5.04E-02 |
| rs4664443 | G | 0.27 | 0.46 | 5.50E-01 | 2.36 | 1.45 | 1.03E-01 | 1.85 | 1.13 | 1.02E-01 |
| rs2284870 | G | 0.51 | 0.48 | 2.91E-01 | 3.77 | 1.52 | 1.37E-02 | 2.44 | 1.19 | 4.08E-02 |
| **rs6432708** | **C** | -0.58 | 0.47 | **2.17E-01** | **-4.98** | **1.47** | **7.69E-04** | **-3.28** | **1.15** | **4.44E-03** |
| rs10930040 | G | 0.92 | 0.50 | 6.87E-02 | 5.01 | 1.59 | 1.65E-03 | 2.61 | 1.24 | 3.56E-02 |
| rs71408196 | I | 1.12 | 0.72 | 1.22E-01 | 5.99 | 2.28 | 8.63E-03 | 3.13 | 1.78 | 7.92E-02 |
| rs12995983 | C | 0.24 | 0.51 | 6.37E-01 | -1.96 | 1.61 | 2.24E-01 | -3.10 | 1.26 | 1.38E-02 |
| rs3788979 | A | 0.53 | 0.76 | 4.91E-01 | 2.91 | 2.41 | 2.28E-01 | 0.68 | 1.88 | 7.16E-01 |
| rs16822665 | T | 0.90 | 0.51 | 7.69E-02 | 5.17 | 1.59 | 1.20E-03 | 2.91 | 1.25 | 1.98E-02 |
| **rs4664446** | **G** | -1.48 | 0.45 | **1.10E-03** | **-6.10** | **1.42** | **1.89E-05** | **-2.62** | **1.12** | **1.91E-02** |
| rs873826 | A | -0.33 | 0.48 | 4.98E-01 | -2.67 | 1.53 | 8.14E-02 | -2.08 | 1.19 | 8.05E-02 |
| rs741529 | A | -0.27 | 0.65 | 6.77E-01 | 4.22 | 2.05 | 3.98E-02 | -0.84 | 1.60 | 5.99E-01 |
| rs35280626 | T | 0.00 | 0.51 | 9.98E-01 | -0.82 | 1.61 | 6.11E-01 | -2.11 | 1.25 | 9.27E-02 |
| rs4436979 | C | -0.13 | 0.46 | 7.83E-01 | -2.03 | 1.44 | 1.60E-01 | -0.48 | 1.13 | 6.71E-01 |
| rs2052352 | T | -0.28 | 0.45 | 5.38E-01 | 0.47 | 1.41 | 7.37E-01 | 1.39 | 1.10 | 2.07E-01 |
| rs13431489 | G | -0.21 | 0.46 | 6.38E-01 | 1.27 | 1.44 | 3.78E-01 | 1.61 | 1.12 | 1.52E-01 |
| rs1990761 | G | -0.21 | 0.49 | 6.77E-01 | -2.02 | 1.56 | 1.95E-01 | -0.12 | 1.22 | 9.21E-01 |

Abbreviations: A, minor allele; BETA, regression coefficient; SE, standard error; $P_{asymp}$, asymptotic P-value for linear regression under the additive model adjusted for age and BMI; SNP's representing peaks of association are highlighted in bold.

**Supplementary Table 5 -** Quantitative trait locus analysis of plasma C-peptide levels at 0min, 30min and 120min during the OGTT with CD26/DPP4 SNPs in NGT subjects.

| | | C-peptide 0min | | | C-peptide 30min | | | C-peptide 120min | | |
|---|---|---|---|---|---|---|---|---|---|---|
| **SNP ID** | **A** | **BETA** | **SE** | **$P_{asymp}$** | **BETA** | **SE** | **$P_{asymp}$** | **BETA** | **SE** | **$P_{asymp.}$** |
| rs10930036 | T | -0.03 | 0.04 | 4.45E-01 | -0.18 | 0.14 | 1.88E-01 | -0.47 | 0.18 | 9.83E-03 |
| rs2052400 | A | -0.10 | 0.04 | 1.71E-02 | -0.26 | 0.13 | 5.36E-02 | -0.54 | 0.17 | 1.84E-03 |
| rs12617656 | C | 0.12 | 0.04 | 5.94E-03 | 0.08 | 0.14 | 6.00E-01 | 0.64 | 0.19 | 7.07E-04 |
| rs2970933 | A | -0.10 | 0.04 | 1.43E-02 | -0.27 | 0.13 | 4.40E-02 | -0.56 | 0.17 | 1.45E-03 |
| rs2287509 | G | -0.10 | 0.04 | 1.70E-02 | -0.29 | 0.14 | 3.01E-02 | -0.53 | 0.18 | 2.47E-03 |
| **rs7565794** | **C** | **0.11** | **0.04** | **1.02E-02** | **0.02** | **0.14** | **9.16E-01** | **0.66** | **0.19** | **4.04E-04** |
| rs2909449 | G | -0.10 | 0.04 | 1.21E-02 | -0.29 | 0.13 | 3.23E-02 | -0.54 | 0.17 | 2.03E-03 |
| rs2909450 | A | 0.00 | 0.05 | 9.18E-01 | 0.09 | 0.15 | 5.57E-01 | 0.02 | 0.19 | 9.03E-01 |
| **rs1014444** | **G** | **0.12** | **0.04** | **4.25E-03** | **0.08** | **0.14** | **5.54E-01** | **0.66** | **0.18** | **2.86E-04** |
| rs1014445 | A | -0.10 | 0.04 | 1.91E-02 | -0.25 | 0.13 | 6.34E-02 | -0.54 | 0.18 | 2.26E-03 |
| rs2268890 | A | -0.10 | 0.04 | 1.30E-02 | -0.27 | 0.13 | 4.38E-02 | -0.55 | 0.17 | 1.48E-03 |
| **rs2300757** | **G** | **0.12** | **0.04** | **5.40E-03** | **0.03** | **0.15** | **8.26E-01** | **0.69** | **0.19** | **2.91E-04** |
| rs12692646 | A | 0.11 | 0.04 | 1.01E-02 | 0.07 | 0.15 | 6.30E-01 | 0.65 | 0.19 | 6.68E-04 |
| rs138687963 | D | -0.01 | 0.05 | 7.94E-01 | 0.07 | 0.15 | 6.41E-01 | 0.04 | 0.19 | 8.23E-01 |
| rs13409045 | T | 0.09 | 0.04 | 2.08E-02 | 0.19 | 0.13 | 1.53E-01 | 0.42 | 0.17 | 1.50E-02 |
| rs2300755 | T | 0.10 | 0.04 | 1.12E-02 | 0.22 | 0.14 | 9.96E-02 | 0.52 | 0.18 | 3.75E-03 |
| rs2268889 | A | 0.11 | 0.04 | 1.08E-02 | 0.11 | 0.14 | 4.38E-01 | 0.56 | 0.19 | 2.80E-03 |
| rs4664443 | G | 0.09 | 0.04 | 1.92E-02 | 0.20 | 0.13 | 1.37E-01 | 0.47 | 0.17 | 7.18E-03 |
| rs2284870 | G | 0.12 | 0.04 | 3.63E-03 | 0.14 | 0.14 | 3.18E-01 | 0.57 | 0.18 | 1.92E-03 |
| rs6432708 | C | -0.11 | 0.04 | 8.85E-03 | -0.28 | 0.14 | 4.29E-02 | -0.54 | 0.18 | 2.44E-03 |
| rs10930040 | G | 0.11 | 0.04 | 1.34E-02 | 0.05 | 0.15 | 7.29E-01 | 0.60 | 0.19 | 1.64E-03 |
| rs71408196 | I | 0.15 | 0.06 | 2.16E-02 | 0.21 | 0.21 | 3.23E-01 | 0.79 | 0.27 | 4.09E-03 |
| rs12995983 | C | -0.09 | 0.04 | 3.80E-02 | -0.21 | 0.15 | 1.50E-01 | -0.49 | 0.19 | 1.08E-02 |
| rs3788979 | A | -0.04 | 0.07 | 5.04E-01 | -0.07 | 0.22 | 7.57E-01 | -0.10 | 0.29 | 7.19E-01 |
| **rs16822665** | **T** | **0.12** | **0.04** | **7.71E-03** | **0.08** | **0.15** | **5.69E-01** | **0.65** | **0.19** | **7.03E-04** |
| rs4664446 | G | -0.08 | 0.04 | 4.27E-02 | -0.07 | 0.13 | 5.74E-01 | -0.40 | 0.17 | 1.87E-02 |
| rs873826 | A | -0.09 | 0.04 | 4.39E-02 | -0.07 | 0.14 | 5.93E-01 | -0.40 | 0.18 | 3.06E-02 |
| rs741529 | A | 0.04 | 0.06 | 4.45E-01 | 0.18 | 0.19 | 3.51E-01 | 0.33 | 0.25 | 1.82E-01 |
| rs35280626 | T | -0.06 | 0.04 | 2.16E-01 | 0.00 | 0.15 | 9.93E-01 | -0.23 | 0.19 | 2.36E-01 |
| rs4436979 | C | 0.02 | 0.04 | 5.60E-01 | -0.08 | 0.13 | 5.56E-01 | -0.15 | 0.17 | 3.81E-01 |
| rs2052352 | T | 0.04 | 0.04 | 3.31E-01 | 0.16 | 0.13 | 2.02E-01 | 0.14 | 0.17 | 3.99E-01 |
| rs13431489 | G | 0.02 | 0.04 | 5.99E-01 | 0.12 | 0.13 | 3.65E-01 | 0.17 | 0.17 | 3.16E-01 |
| rs1990761 | G | -0.02 | 0.04 | 6.32E-01 | 0.07 | 0.14 | 6.42E-01 | -0.13 | 0.19 | 4.71E-01 |

Abbreviations: A, minor allele; BETA, regression coefficient; SE. standard error; $P_{asymp}$, asymptotic P-value for linear regression under the additive model adjusted for age and BMI; SNP's representing peaks of association are highlighted in bold.

**Supplementary Figure 1**

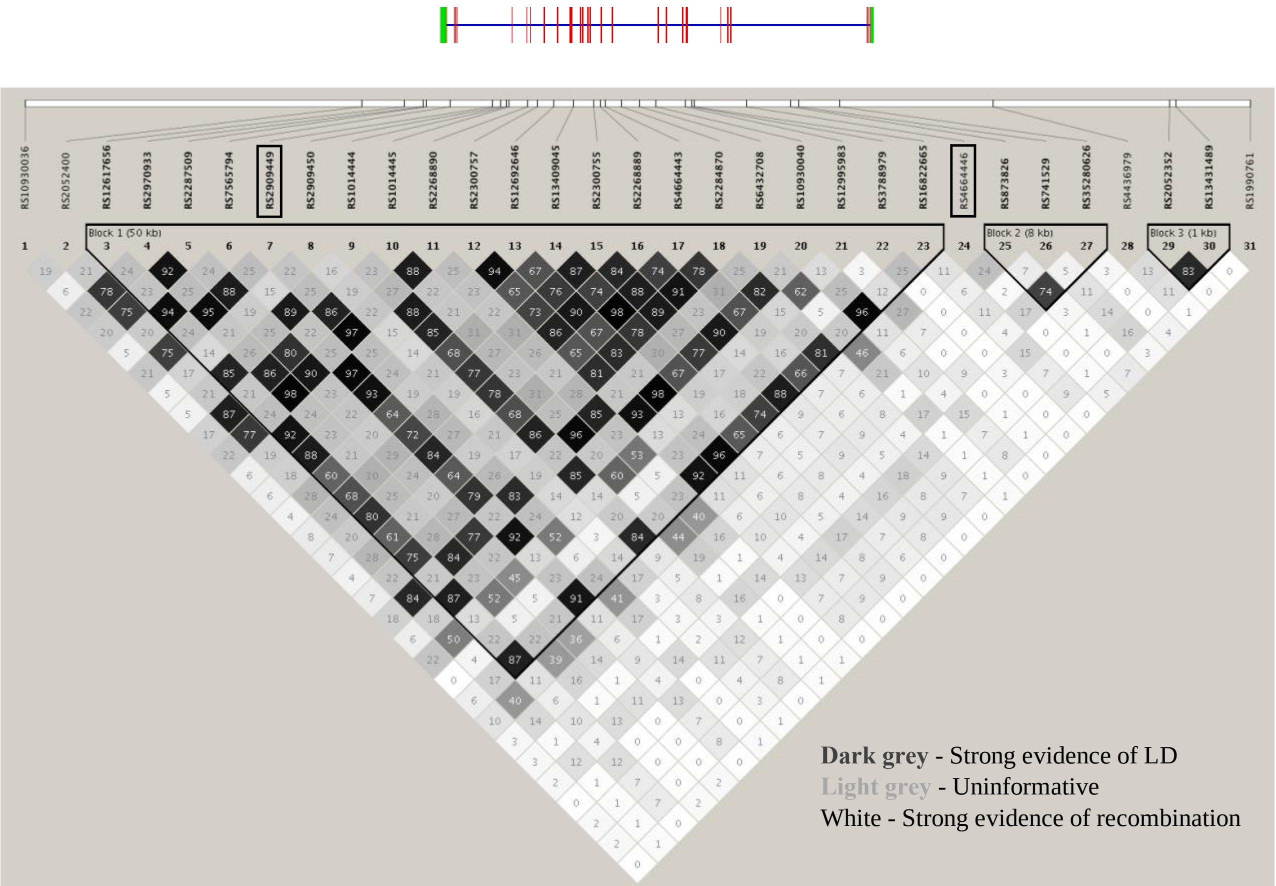


**Supplementary Figure 1.** Linkage disequilibrium (LD) map of the genotyped CD26/DPP4 SNPs with overimposed scaled representation of exon-intron structure of the CD26/DPP4 gene (oriented from right to left). R-square values for pairwise LD were calculated using 969 subjects analyzed in this study. Color coding highlights a large LD block spanning from intron 5 to 25 (Block 1). Indels (rs138687963 and rs71408196) are excluded from the LD map. LD plot was generated by Haploview 4.2. software.

**Supplementary Figure 2**

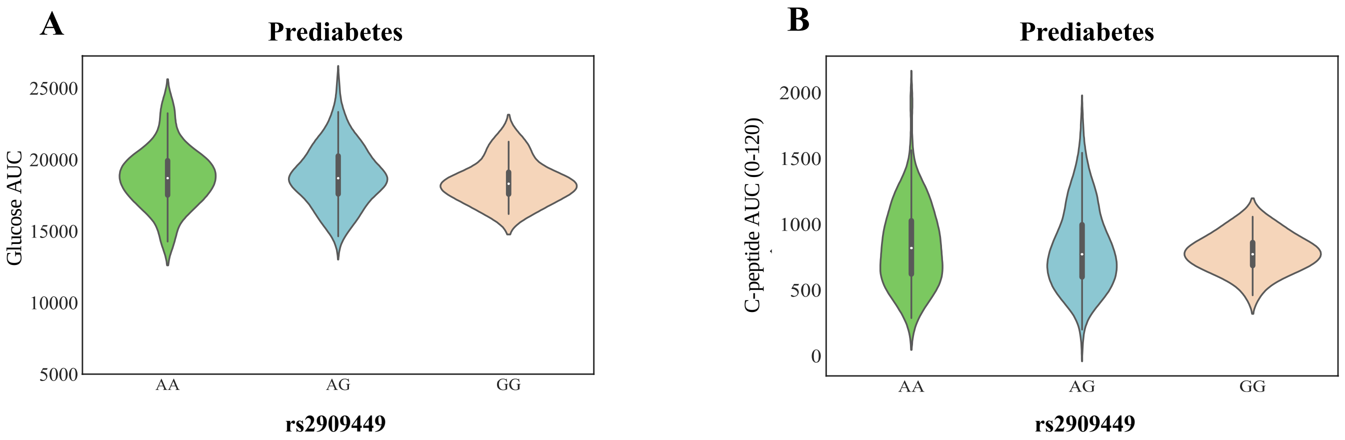


**Supplementary Figure 2.** Violin plots of genotype-class effects of rs2909449 on plasma glucose AUC (A) and C-peptide AUC (0-120) (B) during OGTT (green, ancestral allele homozygotes; blue heterozygotes; pink, minor allele homozygotes). Kruskal-Wallis test with Dunn's correction for multiple comparisons (*P<0.05, **P<0.01 and ***P<0.001, unpaired t-test Mann-Whitney). The plots represent the probability density, the median, the interquartile range and the 95% confidence interval of the phenotypic distributions per genotype class totaling 233 prediabetic subjects.
